# Risk of Long Covid in people infected with SARS-CoV-2 after two doses of a COVID-19 vaccine: community-based, matched cohort study

**DOI:** 10.1101/2022.02.23.22271388

**Authors:** Daniel Ayoubkhani, Matthew L. Bosworth, Sasha King, Koen B. Pouwels, Myer Glickman, Vahé Nafilyan, Francesco Zaccardi, Kamlesh Khunti, Nisreen A. Alwan, A. Sarah Walker

## Abstract

**Background:** It is unclear whether receiving two COVID-19 vaccinations before SARS-CoV-2 infection reduces the risk of developing Long Covid symptoms. We examined whether the likelihood of symptoms 12 weeks after infection differed by vaccination status.

**Methods:** We included COVID-19 Infection Survey participants aged 18-69 years who tested positive for SARS-CoV-2 between 26 April 2020 and 30 November 2021; we excluded participants who, before their first test-confirmed infection, had suspected COVID-19 or Long Covid symptoms, or were single-vaccinated. Participants who were double-vaccinated ≥14 days before infection were 1:1 propensity-score matched, based on socio-demographic characteristics and time from infection to follow-up for Long Covid, to those unvaccinated at time of infection. We estimated adjusted odds ratios (aOR) of Long Covid symptoms ≥12 weeks post-infection, comparing double-vaccinated with unvaccinated (reference group) participants.

**Results:** The study sample comprised 3,090 double-vaccinated participants (mean age 49 years, 54% female, 92% white, median follow-up from infection 96 days) and matched control participants. Long Covid symptoms were reported by 294 double-vaccinated participants (prevalence 9.5%) compared with 452 unvaccinated participants (14.6%), corresponding to an aOR for Long Covid symptoms of 0.59 (95% CI: 0.50 to 0.69). There was no evidence of heterogeneity by adenovirus vector versus messenger ribonucleic acid vaccines (p=0.25).

**Conclusions:** COVID-19 vaccination is associated with reduced risk of Long Covid, emphasising the need for public health initiatives to increase population-level vaccine uptake. Longer follow-up is needed, as is the assessment of further vaccine doses and the Omicron variant.

## Introduction

Long-term symptoms following SARS-CoV-2 infection, often referred to as Long Covid, post-COVID-19 syndrome, post-acute COVID-19 syndrome, post-COVID condition, or post-acute sequelae of SARS-CoV-2 infection, affect approximately 2% of the UK population, with two-thirds of these individuals experiencing functional impairment [1]. COVID-19 vaccines reduce rates of SARS-CoV-2 infection [2] and transmission [3] and therefore Long Covid incidence. However, it is unclear to what extent vaccination reduces the risk of developing Long Covid symptoms following breakthrough infection, with mixed evidence to date [4,5].

To 25 January 2022, 16% of the UK population eligible for a second vaccination were yet to receive it [6], while vaccine coverage was lowest in disadvantaged groups, including ethnic minorities and deprived communities, where rates of infection have been highest [7]. Understanding the role of vaccines in Long Covid may therefore aid public health messaging and facilitate informed decision-making regarding vaccine uptake. We investigated whether infection following two doses of a COVID-19 vaccine is associated with a reduction in Long Covid symptoms after 12 weeks, relative to being unvaccinated when infected.

## Methods

### Study data and design

The main data source was the UK COVID-19 Infection Survey (CIS, ISRCTN21086382, https://www.ndm.ox.ac.uk/covid-19/covid-19-infection-survey/protocol-and-information-sheets), comprising a sample of over half a million participants randomly selected from the UK community population. Ethical approval was obtained from the South Central Berkshire B Research Ethics Committee (20/SC/0195). At enrolment, adult participants provided written consent, including for optional weekly follow-up visits for one month followed by at least 12 monthly visits in the majority.

We included CIS participants aged 18-69 years who tested positive for SARS-CoV-2, either by polymerase chain reaction test using swabs obtained at study visits (58.7% of infections) or any swab test in national testing programmes (self-reported by study participants), between 26 April 2020 and 30 November 2021. We excluded participants who: reported suspected COVID-19 or tested positive for antibodies (in the study or elsewhere) more than two weeks before their first positive swab; reported Long Covid symptoms at any time before their first positive swab; had never responded to the survey question on Long Covid (see ‘Outcome’ below) following its introduction on 3 February 2021; did not have ≥12 weeks of post-infection follow-up by 30 November 2021; or were single-vaccinated when infected.

### Exposure

The exposure of interest was receipt of at least two doses of a COVID-19 vaccine (Oxford/AstraZeneca ChAdOx1 nCoV-19 [AZD1222], Pfizer/BioNTech BNT162b2, or Moderna mRNA-1273) ≥14 days before the first test-confirmed infection. Vaccination status for participants in England was derived from survey data linked to National Immunisation Management System records, with the latter being prioritised where they conflicted with self-reports. Administrative data were not available for participants in Wales, Scotland, and Northern Ireland (13.6%), thus vaccination status was derived solely from self-report.

### Outcome

The primary outcome was Long Covid status according to the survey question: “Would you describe yourself as having ‘Long Covid’, that is, you are still experiencing symptoms more than 4 weeks after you first had COVID-19, that are not explained by something else?” Participants were also asked whether their symptoms limited their ability to undertake daily activities. We considered participants’ first response ≥12 weeks after their first test-confirmed infection (defined as follow-up time).

### Statistical methods

We matched study participants who were double-vaccinated at time of infection to control participants who were unvaccinated when infected and remained so at their first follow-up visit ≥12 weeks later. Double-vaccinated and unvaccinated participants were 1:1 propensity-score matched within calipers of 0.1 points of the propensity score on socio-demographic characteristics: age, sex, ethnicity (white or non-white), country/region of residence, area deprivation quintile group and self-reported, pre-existing health/disability status. Large imbalance after matching was identified by absolute standardized differences >10% [8]. Although a ‘post-treatment’ variable, we also included time from infection to follow-up for Long Covid in the matching set to avoid evaluating Long Covid symptoms in unvaccinated and double-vaccinated participants at different stages of the illness **(Supplementary Figure 1)**. Continuous variables were modelled as restricted cubic splines, with boundary knots at the 10^th^ and 90^th^ percentiles and an internal knot at the median of the distributions.

We estimated adjusted odds ratios (aOR) for Long Covid at ≥12 weeks using logistic regression including all covariates from the matching set, comparing participants who were double-vaccinated to those unvaccinated (reference group) when infected, using robust standard errors to account for matching. We tested for effect modification by time from infection to follow-up for Long Covid, and by adenovirus vector (Oxford/AstraZeneca) versus messenger ribonucleic acid (mRNA; Pfizer/BioNTech or Moderna) vaccines. Statistical analyses were performed using R version 3.6.

## Results

Of 3,333 eligible participants who were double-vaccinated before their first test-confirmed SARS-CoV-2 infection, 3,090 (92.7%) were 1:1 matched to participants who were unvaccinated when infected (from a pool of 9,854 potential control participants). Among double-vaccinated participants, 2,287 (74.0%), 788 (25.5%) and 15 (0.5%) received Oxford/AstraZeneca, Pfizer/BioNTech, and Moderna vaccines, respectively.

Most double-vaccinated participants (3,057, 98.9%) were infected after 17 May 2021, when the Delta variant dominated in the UK, while nearly all unvaccinated participants (3,082, 99.7%) were infected before this date. Median follow-up for Long Covid ≥12 weeks after infection among double-vaccinated and unvaccinated participants was 96 (IQR: 90 to 104) and 98 (89 to 109) days, respectively. After matching, socio-demographic characteristics were generally well balanced for all variables except age (mean 49 versus 47 years for double-vaccinated versus unvaccinated, absolute standardized difference 19.6%) **(Table 1)**.

**Table 1.**
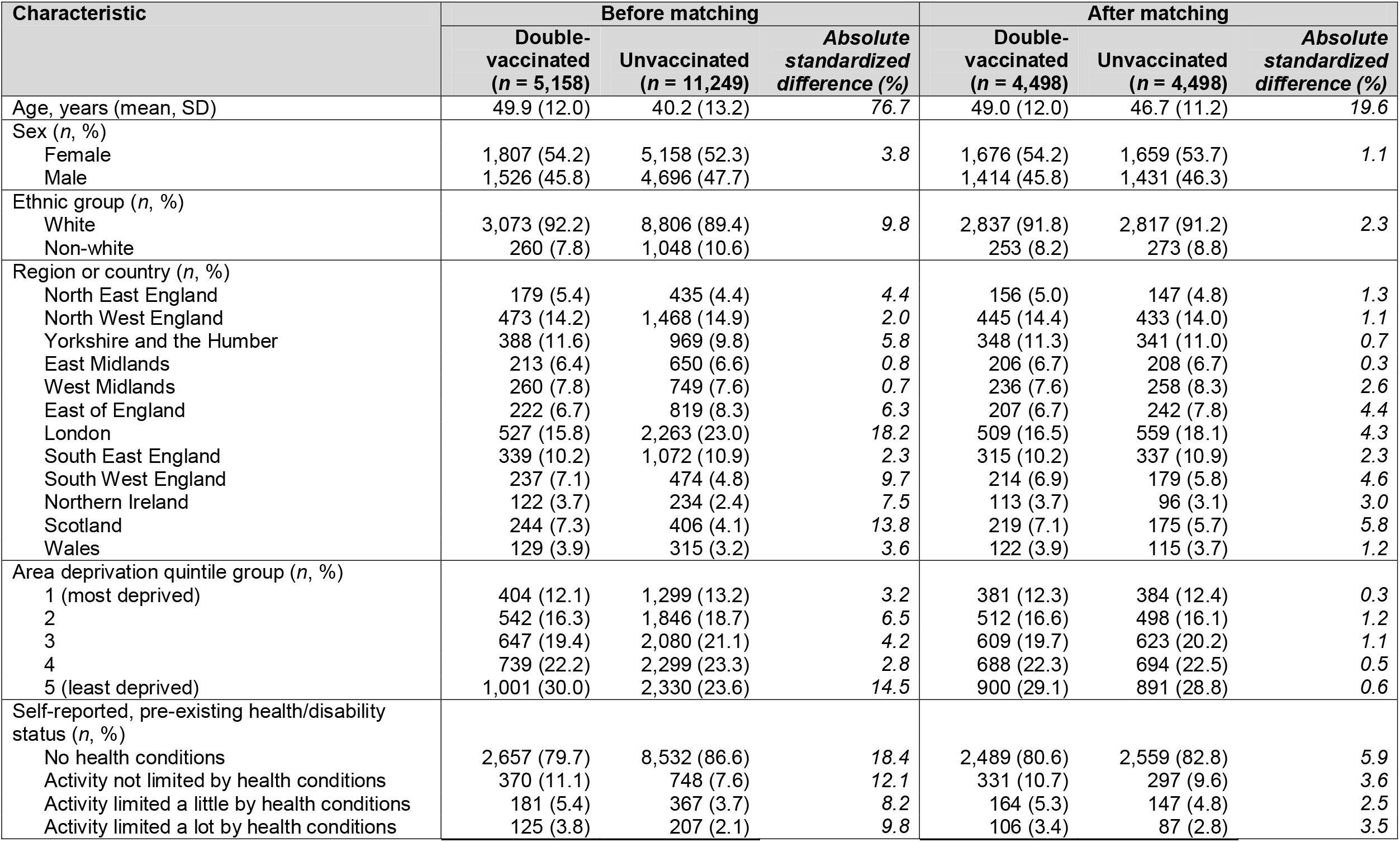
Characteristics of study participants who were double-vaccinated or unvaccinated at the time of infection, before and after matching

At follow-up, Long Covid symptoms of any severity were reported by 294 double-vaccinated participants (prevalence 9.5%; 95% CI: 8.5% to 10.6%) versus 452 unvaccinated participants (14.6%; 13.4% to 15.9%), and activity-limiting symptoms by 170 (5.5%; 4.8% to 6.4%) and 268 (8.7%; 7.7% to 9.7%) participants, respectively.

The aOR were 0.59 (0.50 to 0.69) for Long Covid of any severity and 0.59 (0.48 to 0.73) for activity-limiting symptoms in those infected after double vaccination compared with those who were infected when unvaccinated **(Figure 1)**. There was no evidence of heterogeneity by time from infection to follow-up (p=0.65 for symptoms of any severity; p=0.68 for activity-limiting symptoms), or between participants receiving adenovirus vector or mRNA vaccines (p=0.25 for symptoms of any severity; p=0.35 for activity-limiting symptoms).

**Figure 1.**
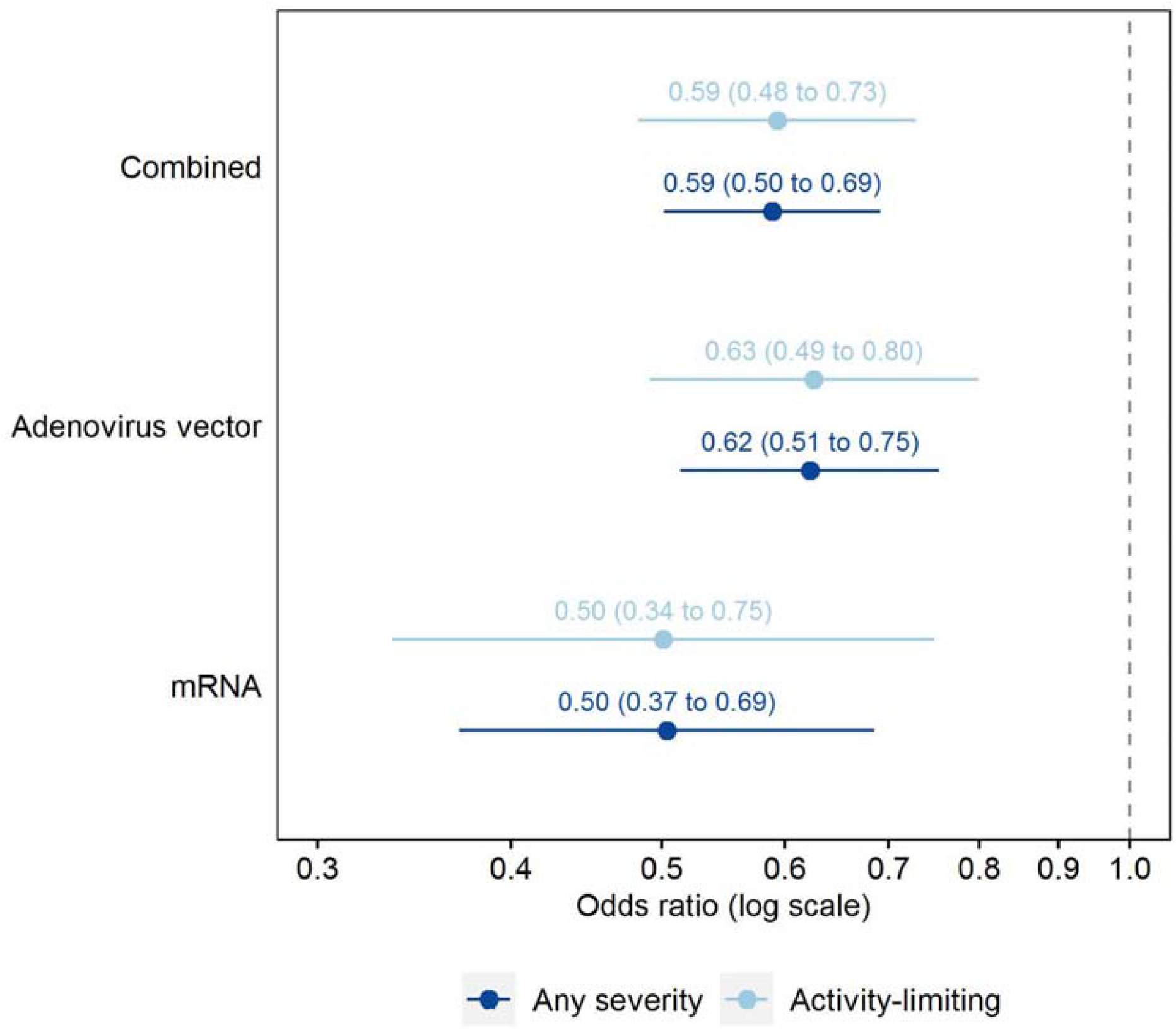
Adjusted odds ratios for Long Covid symptoms ≥12 weeks after first infection, comparing matched study participants who were double-vaccinated or unvaccinated (reference group) before infection Odds ratios adjusted for socio-demographic characteristics (age, sex, white or non-white ethnicity, country/region of residence, area deprivation quintile group, and self-reported, pre-existing health/disability status) and time from infection to follow-up for Long Covid. Confidence intervals are at the 95% level.

Sensitivity analysis demonstrated that the aOR increased when removing time from infection to follow-up for Long Covid from the matching set (to 0.68 [0.56 to 0.81] for the primary outcome), and further increased when it was also omitted from the covariate set in adjusted models (0.73 [0.62 to 0.85]) **(Supplementary Table 1)**. However, the aOR remained below 1 in all analyses.

## Discussion

We found that receiving two COVID-19 vaccinations at least two weeks before SARS-CoV-2 infection was associated with a 41% decrease in the odds of developing Long Covid symptoms 12 weeks later, relative to not being vaccinated when infected. Our results extend those already published, whereby the risk of Long Covid was approximately halved in people who were double-vaccinated when infected compared with those who were unvaccinated, but at four rather than 12 weeks of follow-up [4]. Conclusions based on healthcare records rather than self-report (as in our study) are less clear, with vaccination associated with reduced rates of only specific symptoms [5] and diagnoses [9], though under-presentation, under-diagnosis, and under-recording are all possible [10]. In addition to reducing the risk of Long Covid after breakthrough infection, evidence also suggests that the likelihood and severity of pre-existing Long Covid symptoms may be reduced after vaccination [11,12].

The main study strength is that the CIS comprises a large sample of participants randomly selected from the population to minimise selection bias. Participants are routinely tested for SARS-CoV-2 at follow-up visits, therefore our study includes both asymptomatic and symptomatic infections, as well as self-reported tests. We adjusted for multiple factors related to vaccination uptake [7] and long-term symptoms [13]. However, as with any observational study, some unmeasured confounding may remain. In particular, because the question on Long Covid was not introduced until 3 February 2021, shortly after mass COVID-19 vaccination started in the UK on 8 December 2020, one key limitation is that it was not possible to match double-vaccinated and unvaccinated participants on calendar time of infection **(Supplementary Figure 1)**. Differences in the likelihood of developing Long Covid symptoms between exposure groups may therefore partly reflect changes in the dominant COVID-19 variant or other period effects. Our key exposure was double vaccination, despite third and booster doses now being available, and the study period was before the Omicron variant became widespread. We were not able to investigate participants who were single-vaccinated when infected because nearly all of these received their second dose within the 12-week follow-up period, confounding any relationship between one dose at infection and Long Covid symptoms.

In conclusion, SARS-CoV-2 infection after double vaccination is associated with a reduced risk of developing Long Covid symptoms at 12 weeks compared with infection before vaccination, emphasising the need for public health initiatives to increase population-level vaccine uptake. Studies with longer follow-up are needed to assess the impact of booster doses and the Omicron variant and to evaluate symptom trajectories beyond a single 12-week follow-up visit, particularly given the relapsing nature of Long Covid [14]. Further research into possible biological explanations behind our findings, which may inform therapeutic strategies for Long Covid, is also required.

## Data Availability

De-identified study data are available to accredited researchers in the ONS Secure Research Service (SRS) under part 5, chapter 5 of the Digital Economy Act 2017. For further information about accreditation, contact or visit: https://www.ons.gov.uk/aboutus/whatwedo/statistics/requestingstatistics/approvedresearcherscheme

## Acknowledgements

KK and FZ are supported by the National Institute for Health Research (NIHR) Applied Research Collaboration East Midlands (ARC EM) and the NIHR Leicester Biomedical Research Centre (BRC). KBP and ASW are supported by the NIHR Health Protection Research Unit in Healthcare Associated Infections and Antimicrobial Resistance (NIHR200915), a partnership between the UK Health Security Agency (UKHSA) and the University of Oxford. KBP is also supported by the Huo Family Foundation. ASW is also supported by the NIHR Oxford Biomedical Research Centre and is an NIHR Senior Investigator. NAA has lived experience of Long Covid and is a co-investigator on the NIHR-funded STIMULATE-ICP study. The views expressed are those of the authors and are not necessarily those of the National Health Service, the NIHR, the Department of Health and Social Care, or the UK Health Security Agency. For the purpose of open access, the authors have applied a Creative Commons Attribution (CC BY) licence to any Author Accepted Manuscript version arising.

## Footnotes

### Data availability

De-identified study data are available to accredited researchers in the ONS Secure Research Service (SRS) under part 5, chapter 5 of the Digital Economy Act 2017. For further information about accreditation, contact or visit: https://www.ons.gov.uk/aboutus/whatwedo/statistics/requestingstatistics/approvedresearcher scheme

### Author contributions

All authors contributed to conceptualising and designing the study. DA, MLB and SK prepared the study data and performed the statistical analysis. All authors contributed to interpretation of the results. DA, MLB and SK were responsible for the first draft of the manuscript. All authors contributed to critical revision of the manuscript. All authors approved the final manuscript.

### Ethical approval

Ethical approval for this study was obtained from the National Statistician’s Data Ethics Advisory Committee (NSDEC(20)12). The CIS received ethical approval from the South Central Berkshire B Research Ethics Committee (20/SC/0195).

---

**Supplementary Table 1.** Adjusted odds ratios for the main analysis (Approach 1) and sensitivity analyses whereby follow-up time from infection to follow-up for Long Covid ≥12 weeks later was removed from the matching set (Approach 2) and the adjusted models (Approach 3)

| Outcome | Vaccine type | Approach 1 | Approach 2 | Approach 3 |
| --- | --- | --- | --- | --- |
| Long Covid of any severity | Combined | 0.59 (0.50 to 0.69) | 0.68 (0.56 to 0.81) | 0.73 (0.62 to 0.85) |
|  | Adenovirus vector | 0.62 (0.51 to 0.75) | 0.74 (0.61 to 0.91) | 0.80 (0.67 to 0.96) |
|  | mRNA | 0.50 (0.37 to 0.69) | 0.52 (0.38 to 0.72) | 0.56 (0.41 to 0.76) |
| Activity-limiting Long Covid | Combined | 0.59 (0.48 to 0.73) | 0.65 (0.51 to 0.82) | 0.69 (0.56 to 0.84) |
|  | Adenovirus vector | 0.63 (0.49 to 0.80) | 0.73 (0.56 to 0.95) | 0.77 (0.61 to 0.97) |
|  | mRNA | 0.50 (0.34 to 0.75) | 0.47 (0.31 to 0.72) | 0.50 (0.34 to 0.74) |

**Supplementary Figure 1.**
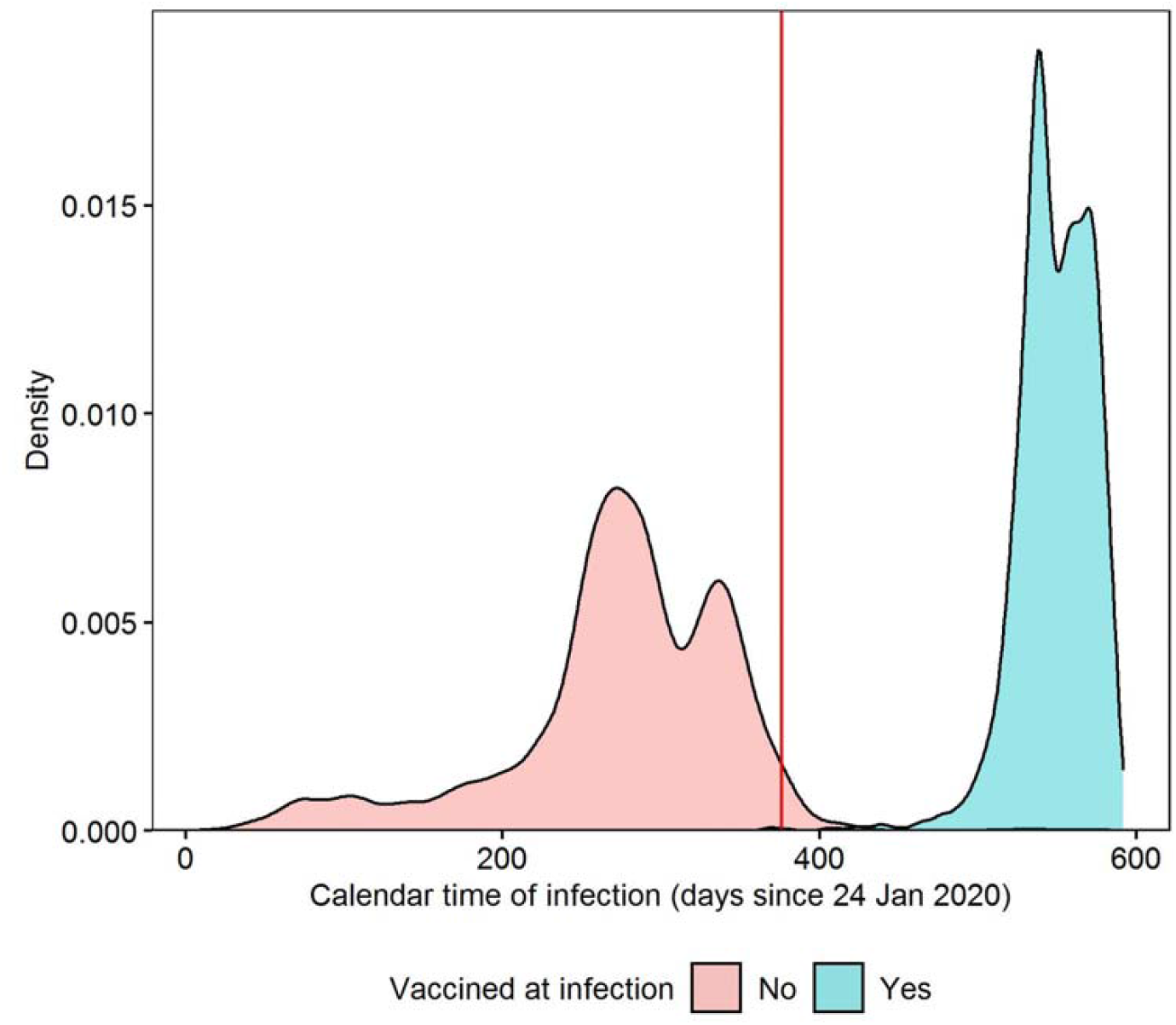
Density plot of calendar time of first infection, stratified by whether study participants were double-vaccinated ≥14 days before infection; the red line indicates the introduction of the survey question on Long Covid on 3 February 2021 Calendar time of infection calculated as the number of days from 24 January 2020, when the first COVID-19 case was reported in the UK. Density estimated from 3,333 double-vaccinated participants and 9,854 unvaccinated participants before matching.

Supplementary Figure 1 demonstrates that there was almost no common support in the distribution of calendar time of infection stratified by vaccination status when infected. This means that it was not possible to match double-vaccinated and unvaccinated participants on calendar time of infection.

Furthermore, the position of the red vertical line in Supplementary Figure 1, denoting the introduction of the survey question on Long Covid on 3 February 2021, illustrates why time from infection to follow-up for Long Covid ≥12 weeks later tended to be longer for unvaccinated than double-vaccinated participants. It was therefore necessary to match on this duration, to avoid evaluating Long Covid symptoms in unvaccinated and double-vaccinated participants at different stages of the illness as it progresses.

